# Determinants of specificity and end-user acceptability of an IP-10-based point-of-care triage test for antiretroviral therapy monitoring in Mozambique

**DOI:** 10.64898/2026.05.22.26353111

**Authors:** Anna Saura-Lázaro, Dulce Adolfo Bila, Erika van den Bogaart, Hanlie Myburgh, Madina Fischer-Cunhete, Paula Vaz, René Paulussen, Lario Viljoen, Tobias F. Rinke de Wit, Denise Naniche

## Abstract

**Introduction:** Viral load (VL) monitoring is the gold standard for antiretroviral therapy (ART) monitoring. Still, due to limited funds and infrastructure, many people living with HIV (PLHIV) in low– and middle-income countries do not receive timely VL testing. We evaluated the clinical performance and end-user acceptability of a prototype interferon gamma-induced protein 10 (IP-10) point-of-care (POC) test as a rule-out triage tool to identify individuals unlikely to have unsuppressed VL in PLHIV in Mozambique.

**Methods:** A mixed-methods study was conducted between November 2023 and November 2024 at two primary healthcare facilities in Maputo Province. We enrolled 1,057 PLHIV on ART from stable and specialized risk clinics. Clinical performance of the IP-10 POC test (index test) was compared against plasma HIV VL (reference test; unsuppressed defined as >1000 copies/mL). Socio-demographic and clinical predictors of false-positive results were identified using multivariable logistic regression. Immediate acceptability was assessed through exit interviews on a subset of 43 PLHIV.

**Results:** Among participants (71.7% female; median age 41.4 years), 12.0% had unsuppressed VL. The IP-10 POC test demonstrated high sensitivity (90.6%) and moderate specificity (35.6%). Specificity was higher in clinics treating stable patients (44.5% 95%CI: 39.7−49.3) compared to specialized risk clinics (26.5% 95%CI: 21.1−28.9). The proportion of false-positive results was also higher in patients attending specialized risk clinics. Independent predictors of false positivity included enrolment in a “one-stop” TB/HIV clinic (aOR=2.99 95%CI: 1.09−8.15), cotrimoxazole use (aOR=2.16, 95% CI: 1.13−4.13), and obesity (aOR=3.47 95%CI: 1.74−6.93). Acceptability was high: 70% of participants appreciated the test’s simplicity and rapid results, and 95.3% expressed interest in future testing. Most patients preferred finger-prick collection over venous draws.

**Conclusions:** The IP-10 POC test is a highly sensitive triage tool, demonstrating superior performance among stable PLHIV enrolled in differentiated service delivery models like six-month multi-month dispensing. While factors associated with co-infections can reduce specificity, the test’s high acceptability and potential to reduce confirmatory VL test demand suggests it could serve as a viable triage strategy for optimizing resources particularly in stable care pathways with a lower prevalence of inflammatory comorbidities. This could enable health systems to reallocate intensive monitoring toward higher-risk populations.

## Introduction

The World Health Organisation (WHO) recommends individual laboratory nucleic acid amplification testing (NAAT)-based viral load (VL) testing on plasma as the gold standard for monitoring antiretroviral therapy (ART) effectiveness in people living with HIV (PLHIV). VL monitoring is essential to detect virologic failure early, prevent treatment failure, and reduce HIV incidence through treatment as prevention. However, it faces significant challenges in many low– and middle-income countries: limited laboratory infrastructure, cold chain needs, and logistical constraints which lead to highly variable coverage (1). While the use of dried blood spots simplifies sample collection, centralisation, high costs, and delayed results remain major barriers (2). Moreover, most routine VL tests yield undetectable results (3–5), highlighting inefficiencies and the need for more cost-effective ART monitoring strategies. The recent reduction of U.S. funding for HIV programs adds further pressure on already strained healthcare systems to optimize resources while maintaining clinical standards (6).

Point-of-care (POC) assays for HIV VL measurement offer a compelling alternative to centralised VL testing for monitoring ART among PLHIV as they can overcome challenges related to costs, specimen transport and result delivery (7,8). The currently available NAAT-based HIV VL POC technologies are suboptimal, still implying costs, equipment maintenance, sample preparation, and may not effectively reach the most vulnerable populations (9,10).

The current WHO ART monitoring algorithm recommends VL testing six and twelve months after ART initiation, and annually thereafter for individuals established on ART. If the first VL is detectable (>50 copies/mL), three-monthly visits for enhanced adherence counselling (EAC) are advised, monitored by repeat VL testing after three months (11).

To address the challenges associated with NAAT, Mondial Diagnostics, a private non-profit organisation in the Netherlands, developed a prototype lateral flow POC test that measures interferon gamma-induced protein 10 (IP-10), a chemokine shown to strongly correlate with HIV VL (12,13). A clinical performance study conducted in South Africa using the IP-10 POC test as a triage tool to identify PLHIV on ART with unsuppressed HIV VL (>1000 copies/mL), demonstrated high sensitivity and moderate specificity (14). The study also showed the test’s potential cost-effectiveness as a triage tool, reducing by up to 50% the number of VL tests required for ART monitoring; VL testing would only be required for PLHIV who test positive for IP-10. This IP-10 POC triage test does not require specialised equipment, is inexpensive, and potentially amenable to decentralised health facility and community-based testing. However, as a non-specific inflammatory biomarker, the specificity of IP-10 for unsuppressed HIV VL could be impacted by comorbidities.

Implementation of an IP-10 POC triage strategy could improve ART monitoring by reducing the need for VL testing among individuals with negative IP-10 results. This approach would enable same-day ART monitoring results for most patients, reduce unnecessary VL testing and follow-up visits, improve retention in care, and lower health-system costs. Particularly suitable in low-resource settings with a high HIV burden that have reached 95-95-95 testing, treatment and viral load suppression targets. such a test would optimize resource allocation and strengthen differentiated HIV service delivery (DSD) (15). To inform real-world implementation of the IP-10 POC triage test, we assessed factors associated with IP-10 test specificity and explored its acceptability among end users in Mozambique.

## Methods

### Study design, setting, and population

We conducted a mixed-method study in Southern Mozambique integrating a clinical performance evaluation and a nested qualitative acceptability component. The study was performed in the Maputo province between November 2023-November 2024 at a rural and an urban primary healthcare facility in the Magude and Matola districts respectively. (Details in Appendix 1 – supplementary methods). In 2023, the Maputo province (excluding Maputo city), and Gaza province, harbored the highest HIV prevalence in the country, ranging from 22.9% to 24.4%, with higher rates observed in rural areas (16).

PLHIV aged ≥18 years on ART who attended scheduled clinic visits for VL testing or had a VL result available within 30 days were pre-screened for study eligibility. PLHIV attending routine three-month VL follow-up visits after a previous unsuppressed VL result were excluded to allow assessment of the acceptability of implementing the IP-10 POC test from the beginning of the ART monitoring algorithm. We did not apply any restriction regarding time on ART, treatment adherence, or clinical symptoms in order to evaluate the test under real-world conditions and capture data on symptoms or comorbidities potentially associated with elevated IP-10 levels that could reduce the test’s specificity.

For the clinical performance evaluation, assuming a 15% prevalence of unsuppressed VL and based on the prior observed sensitivity (91%) and specificity (44%) of the IP-10 POC test (14), a sample size of 126 PLHIV with unsuppressed VL and a minimum of 380 PLHIV with suppressed VL was calculated to provide a significance level of 0.05 and a confidence interval (CI) half-width of 0.05. To ensure the number of PLHIV with unsuppressed VL and reach 90% statistical power, recruitment from stable patient clinics was enriched with PLHIV attending specialized risk clinics (details in Supplementary materials Annex I).

After reaching nearly 50% of the sample size of PLHIV with unsuppressed VL (52 participants with unsuppressed VL) and 519 with suppressed VL, an interim analysis was conducted establishing the optimal IP-10 POC triage test threshold to be used for the nested qualitative study.

### Study visit procedures

The study included two visits coinciding with scheduled clinic visits per national ART monitoring guidelines, with the second visit occurring approximately 90 days after the first and conducted only for participants with unsuppressed VL. After obtaining informed consent, participants underwent finger-prick capillary blood collection for the IP-10 POC test (index test) and venous phlebotomy performed by a trained nurse. Additionally, a research assistant collected socio-demographic and clinical data via an electronic questionnaire. The IP-10 POC testing was conducted using capillary plasma separated on-site with a portable mini-centrifuge, and results were read using a portable Cube Reader (Chembio Diagnostics) which provides a numerical readout in arbitrary units proportional to IP-10 concentration. The procedure to perform the IP-10 POC test is described elsewhere (10). IP-10 POC test results were disclosed to participants in the nested qualitative study once the optimal threshold for the test was determined. Plasma was recovered from venous blood samples and cryopreserved within six hours of collection for HIV VL determinations (reference test). In accordance with national guidelines, within 15–30 days, VL results were delivered by national HIV program personnel to participants with detectable VL (≥50 copies/mL). IP-10 testing was not used for clinical management which followed current ART monitoring guidelines, including EAC and a 90-day follow-up visit.

### Definitions

Unsuppressed VL was defined as a plasma VL result >1,000 copies/mL. Low level viremia (LLV) was defined as a VL result between 50-1,000 copies/mL

### Statistical analysis

Proportions for categorical variables and the median and interquartile range (IQR) for continuous variables were calculated and compared using χ2 or Fisher’s exact test, and the non-parametric Mann−Whitney U test, respectively, stratified by stable and specialized risk clinic participants.

Receiver operating characteristic analyses were conducted to obtain the area under the curve (AUC), as well as sensitivity and specificity for different IP-10 POC test cut-off values. Optimal thresholds for screening unsuppressed VL were selected using the predefined criterion of prioritizing the first value of sensitivity above 90%, as the IP-10 POC test was validated as a triage test which aims to minimize false negatives.

Logistic regression with penalized likelihood was used to identify predictors of false-positive IP-10 test results among socio-demographic and clinical variables. This analysis was restricted to individuals with a VL≤1,000 copies/mL, including both individuals with a false positive and a true negative result for the IP-10 POC test. Variables with p-values <0.20 in bivariate analyses (level of viremia was fixed) were included in multivariable models, followed by backward stepwise selection where variables with p-values <0.05 entered the model and those with p-values <0.10 were retained. Stata version 17 was used for the analyses.

### Assessing immediate acceptability: sampling and recruitment

After establishing the IP-10 positivity threshold, enrolled participants were classified either as IP-10 POC test positive or negative and were subsequently invited to join a nested adaptive open cohort involving three rounds of qualitative data collection to assess the acceptability of implementing the test. Analysis of these data are reported elsewhere (17).

In this manuscript, we present the results on the immediate acceptability of the IP-10 POC triage test among PLHIV on ART, assessed through exit interviews conducted with a subset of participants from the clinical performance study. We aimed to purposely sample 48 PLHIV on ART for diversity in age, sex, and IP-10 POC result, enrolled in care in stable-patient or specialized risk clinics. We also included sampling from youth clinics to include information from younger PLHIV. Two local research assistants, trained in study procedures and qualitative research methods, used a semi-structured discussion guide to explore immediate perceptions and experiences of the IP-10 POC triage test (Supplementary materials: Appendix I). Interviews were conducted in participants’ preferred language, between 15-20 minutes in length, and audio recorded.

### Qualitative data analysis

Audio recordings of exit interviews were transcribed and translated in summary format into an online data capture form. Summaries captured key themes, illustrative quotations, and relevant participant characteristics. Transcript summaries were coded in <u>ATLAS.ti</u> and grouped, quantifying re-occurring sentiments and themes describing participants’ perceptions and experiences of the IP-10 POC test, and across stable and specialized risk clinics.

### Ethical considerations

The study protocol was approved by the Mozambican National Health Institute ethics board (294/CNBS/23) and regulatory agency (7401/380/ANARME). All participants completed informed consent.

## Results

### Participant characteristics

We enrolled 1,057 participants with a median age of 41.4 years (IQR: 34.5-48.8), of whom 71.7% (n=751) were female (Table 1). More than half (56.9%, n=601) were enrolled from specialized risk clinics. The median time since ART initiation was 6.0 years (IQR 0.5-9.9), and over 96% (n=1020) were on first-line dolutegravir plus two nucleoside reverse transcriptase inhibitors at the time of the study. Over 97% (n=1032) were enrolled in a DSD modality of care which varied by clinic type, with stable clinic participants more frequently enrolled in multi-month dispensing models, while specialized risk clinic participants were more commonly managed in one-stop TB/HIV services.

**Table 1.** Participant socio-demographic and clinical factors according to clinic type (stable, specialized risk). All values are presented as n (%).

|  | Total (n=1057) | Stable clinic (n=446)<sup>a</sup> | Specialized risk clinic (n=601)<sup>a</sup> | p-value |
| --- | --- | --- | --- | --- |
| Socio-demographics |  |  |  |  |
| Biological sex (female) | 751 (71.71) | 355 (79.60) | 396 (65.72) | <0.001 |
| Age (years)<sup>b</sup> | 41.43 (34.47-48.79) | 44.41 (38.21-50.92) | 40.20 (32.64-47.47) | <0.001 |
| Employment |  |  |  |  |
| Employed | 360 (34.06) | 122 (27.35) | 233 (38.77) | <0.001 |
| Part-time | 249 (23.56) | 103 (23.09) | 146 (24.29) |  |
| Not employed | 448 (42.38) | 221 (49.55) | 222 (36.04) |  |
| Marital status |  |  |  |  |
| Married<sup>c</sup> | 568 (53.74) | 227 (50.90) | 340 (56.57) | 0.290 |
| Single | 366 (34.63) | 162 (36.32) | 195 (32.45) |  |
| Divorced | 9 (0.85) | 5 (1.12) | 4 (0.67) |  |
| Widowed | 114 (10.79) | 52 (11.66) | 62 (10.32) |  |
| Educational level |  |  |  |  |
| No primary school | 166 (15.70) | 84 (18.83) | 82 (13.64) | <0.001 |
| Primary school | 545 (51.56) | 244 (54.71) | 300 (49.92) |  |
| Secondary school | 321 (30.37) | 113 (25.34) | 201 (33.44) |  |
| Post-secondary school or university | 25 (2.37) | 5 (1.12) | 18 (3.00) |  |
| Signs and symptoms at study visit 1 |  |  |  |  |
| BMI<sup>d</sup> |  |  |  |  |
| Normal weight | 529 (50.05) | 216 (48.43) | 306 (50.92) | 0.581 |
| Underweight | 70 (6.62) | 29 (6.50) | 39 (6.49) |  |
| Overweight | 287 (27.15) | 120 (26.91) | 166 (27.62) |  |
| Obesity | 171 (16.18) | 81 (18.16) | 90 (14.98) |  |
| Temperature (Celsius)<sup>b</sup> | 36.2 (36-36.4) | 36.1 (36.00-36.40) | 36.3 (36.00-36.50) | <0.001 |
| Blood pressure (n=1006)<sup>e</sup> |  |  |  |  |
| Normal | 327 (32.50) | 123 (31.06) | 197 (32.83) | 0.558 |
| Elevated | 679 (67.50) | 273(68.94) | 403 (67.17) |  |
| Cough |  |  |  |  |
| No | 995 (94.13) | 431 (96.64) | 554 (92.18) | 0.008 |
| Yes (cough for 2 weeks or less) | 43 (4.07) | 9 (2.02) | 34 (5.66) |  |
| Yes (cough for over 2 weeks) | 19 (1.80) | 6(1.35) | 13 (2.16) |  |
| Sore throat | 29 (2.74) | 10 (2.24) | 19 (3.16) | 0.370 |
| Shortness of breath or difficulty breathing | 13 (1.23) | 2 (0.45) | 11 (1.83) | 0.051 |
| Muscle pain | 59 (5.58) | 24 (5.38) | 35 (5.82) | 0.759 |
| Tiredness | 50 (4.73) | 13 (2.91) | 36 (5.99) | 0.020 |
| Headache | 72 (6.81) | 22 (4.93) | 50 (8.32) | 0.032 |
| Diarrhoea | 5 (0.47) | 1 (0.22) | 4 (0.67) | 0.401 |
| Vomiting | 1 (0.09) | 0 | 1 (0.17) | 1.000 |
| Skin eruption | 21 (1.99) | 5 (1.12) | 16 (2.66) | 0.117 |
| Signs and symptoms in the previous 4 weeks |  |  |  |  |
| Fever | 124 (11.73) | 35 (7.85) | 89 (14.81) | **0.001** |
| Sore throat | 48 (4.54) | 8 (1.79) | 37 (6.16) | **0.001** |
| Shortness of breath or difficulty breathing | 31 (2.93) | 9 (2.02) | 21 (3.49) | 0.221 |
| Muscle pain | 77 (7.28) | 16 (3.59) | 61 (10.15) | **<0.001** |
| Tiredness | 82 (7.76) | 28 (6.28) | 53 (8.82) | 0.143 |
| Headache | 138 (13.06) | 34 (7.62) | 103 (17.14) | **<0.001** |
| Diarrhoea | 27 (2.55) | 4 (0.90) | 23 (3.83) | **0.003** |
| Vomiting | 11 (1.04) | 2 (0.45) | 9 (1.50) | 0.130 |
| Skin eruption | 38 (3.60) | 10 (2.24) | 28 (4.66) | **0.039** |
| Night sweats | 14 (1.32) | 6 (1.35) | 8 (1.33) | 0.665 |
| **Comorbidities** |  |  |  |  |
| Diabetes | 1 (0.09) | 0 | 1 (0.17) | 1.000 |
| Hypertension | 120 (11.35) | 49 (10.99) | 71 (11.81) | 0.840 |
| Epilepsy | 7 (0.66) | 0 | 7 (1.16) | **0.033** |
| Past tuberculosis<sup>f</sup> | 49 (4.64) | 20 (4.48) | 28 (4.66) | 0.323 |
| Preventive tuberculosis treatment in the last 12 months (isoniazid) | 235 (22.23) | 20 (4.48) | 214 (35.61) | **<0.001** |
| Cotrimoxazole in the last 12 months | 105 (9.93) | 6 (1.35) | 98 (16.31) | **<0.001** |
| Previous confirmed COVID-19 in the last 3 months | 12 (1.14) | 0 | 12 (2.00) | **0.005** |
| COVID-19 vaccination in the last 6 months |  |  |  |  |
| 1 dose | 224 (21.19) | 110 (24.66) | 113 (18.80) | **<0.001** |
| More than 1 dose | 521 (49.29) | 292 (65.47) | 226 (37.60) |  |
| Diagnosis or suspicion of malaria | 25 (2.37) | 5 (1.12) | 20 (3.33) | **0.017** |
| **HIV clinical history** |  |  |  |  |
| Years since HIV diagnosis (n=1035)<sup>b</sup> | 6.54 (0.64-11.33) | 9.37 (6.24-13.01) | 2.03 (0.50-8.71) | **<0.001** |
| Years since ART initiation (n=1045)<sup>b</sup> | 5.97 (0.54-9.94) | 8.19 (5.76-11.42) | 1.89 (0.50-7.86) | **<0.001** |
| WHO stage at ART initiation |  |  |  |  |
| I or II | 890 (84.20) | 365 (81.84) | 518 (86.19) | **0.048** |
| III or IV | 164 (15.52) | 78 (17.490) | 83 (13.81) |  |
| Unknown | 3 (0.28) | 3 (0.67) | 0 |  |
| WHO stage at study visit 1 |  |  |  |  |
| I or II | 863 (81.65) | 355 (79.60) | 501 (83.368) | **0.047** |
| III or IV | 194 (18.35) | 91 (20.40) | 100 (16.64) |  |
| ART regimen at ART initiation |  |  |  |  |
| DTG+2NRTI | 507 (47.97) | 116 (26.01) | 386 (64.23) | **<0.001** |
| NNRTI+2NRTI | 527 (49.86) | 312 (69.96) | 210 (34.94) |  |
| Other | 23 (2.18) | 18 (4.04) | 5 (0.83) |  |
| Current ART regimen |  |  |  |  |
| DTG+2NRTI | 1020 (96.50) | 433 (97.09) | 577 (96.01) | 0.536 |
| (NNRTI or PI) +2NRTI | 32 (3.03) | 10 (2.24) | 22 (3.66) |  |
| Other | 5 (0.47) | 3 (0.67) | 2 (0.33) |  |
| Undetectable VL (at least once) |  |  |  |  |
| Yes | 550 (52.03) | 407 (91.26) | 137 (22.80) | **<0.001** |
| Unknown | 338 (31.98) | 7 (1.57) | 329 (54.74) |  |
| Last VL value (n=738)<sup>b</sup> | 0 (0-1930) | 0 (0-0) | 4880 (500-25000) | **<0.001** |
| Differentiated service delivery model |  |  |  |  |
| **Not enrolled in any model** | 25 (2.37) | 9 (2.02) | 16 (2.66) | **<0.001** |
| **3MMD** | 506 (47.87) | 235 (52.69) | 265 (44.09) |  |
| **6MMD** | 183 (17.31) | 179 (40.13) | 4 (0.67) |  |
| **One-stop TB/HIV clinic** | 306 (29.23) | 1 (0.22) | 305 (50.75) |  |
| **Community-based adherence club or other** | 34 (3.22) | 22 (4.93) | 11 (1.83) |  |
| **Missed ART during the last month<sup>g</sup>** |  |  |  |  |
| **None** | 699 (66.13) | 375 (84.08) | 320 (53.24) | **<0.001** |
| **Any missed dose** | 359 (33.87) | 71 (15.92) | 281 (46.76) |  |
| **ART interruption (at least once) since ART initiation** | 99 (9.37) | 3 (0.67) | 96 (15.97) | **<0.001** |
| **VL and IP-10 at study visit 1** |  |  |  |  |
| **VL (n=1051)** |  |  |  |  |
| **Undetectable (<50 copies/mL)** | 829 (78.43) | 417 (93.60) | 405 (68.07) | **<0.001** |
| **Low level viremia (50-1000 copies/mL)** | 95 (8.99) | 17 (3.81) | 78 (13.11) |  |
| **Unsuppressed (>1000 copies/mL)** | 127 (12.02) | 12 (2.69) | 112 (18.82) |  |
| **IP-10 POC test reading value (arbitrary units.)<sup>b</sup>** | 8.1 (6.4-11.1) | 7.1 (5.6-9.3) | 9.2 (7.1-12.6) | **<0.001** |

<sup>a</sup>Ten participants from other consultations are not shown in the table when stratifying by clinic type
<sup>b</sup>Median (Interquartile range)
<sup>c</sup>This category includes married participants, civil union or legal partnership
<sup>d</sup>BMI categories: normal weight: 18.5–24.9 kg/m<sup>2</sup>; underweight: <18.5 kg/m<sup>2</sup>; overweight: 25–29.9 kg/m<sup>2</sup>; obesity: ≥30 kg/m<sup>2</sup>
<sup>e</sup>Blood pressure categories: normal: systolic <120 mmHg and diastolic <80 mmHg; elevated: systolic 120-139 mmHg and/or diastolic 80-89 mmHg, or systolic 140-159 mmHg and/or diastolic 90-99 mmHg, or systolic 160-179 mmHg and/or diastolic 100-109 mmHg, or systolic ≥180 mmHg and/or diastolic ≥110 mmHg
<sup>f</sup>Tuberculosis site includes pulmonary, extrapulmonary, and unknown site
<sup>g</sup>This category includes at least one ART dose missed in the month.
For variables with missing values, the total number of observations included is indicated next to the variable name
p < 0.05 indicates statistical significance and are highlighted in bold
Current tuberculosis was also assessed but no cases were reported
**Abbreviations:** ART: antiretroviral therapy, BMI: body mass index, DTG: dolutegravir, MMD: multi-month dispensing, NNRTI: non-nucleoside reverse transcriptase inhibitor, NRTI: nucleoside reverse transcriptase inhibitor, PI: Protease Inhibitor, POC: point-of-care, VL: viral load, WHO: World Health Organisation

A total of 12.0% (n=127) had unsuppressed VL and 9.0% (n=95) had LLV (Table 1). The proportion of individuals with either unsuppressed VL or LLV were both significantly higher in those attending the specialized risk clinic as compared to the stable patient clinic (18.8% vs 2.7% and 13.1% vs 3.8%, respectively, p-value <0.001). Overall, 9.4% (n=99) of participants had a history of treatment interruptions. Of these, 97.0% (n=96) were from specialized risk clinics. In addition, 15.9% (n=71) participants from stable patient clinics and 46.8% (n=281) participants from specialized risk clinics responded that they had missed at least one dose during the last month.

### Clinical performance

Considering the entire participant population, the IP-10 POC test showed an estimated AUC of 0.73 (95%CI: 0.68−0.77) on capillary plasma. Using an optimal threshold IP-10 test reading value of 6.9 on the Cube Reader, according to the predefined criterion described in the methods section, the IP-10 POC test identified unsuppressed VL with a sensitivity of 90.6% (95% CI: 84.1−95.0) and a specificity of 35.3% (95% CI: 30.7−36.9).

Considering that 88.2% (n=112) cases of unsuppressed VL were enrolled from the specialized risk clinics, we assessed the performance according to type of clinic. In the stable patient clinic, the AUC was estimated at 0.71 (95% CI 0.56−0.85). Using an optimal threshold test reading value of 6.7 according to the predefined criterion, the IP-10 POC test identified unsuppressed VL with a sensitivity of 91.7% (95% CI: 61.7−99.8) and a specificity of 44.5% (95% CI: 39.7−49.3 (Table 2). On the other hand, in the specialized risk clinic, AUC was estimated at 0.68 (95% CI 0.62−0.74) and using an optimal threshold test reading value of 7.0, according to the predefined criterion, the IP-10 POC test identified unsuppressed VL with a sensitivity of 90.2% (95% CI: 84.2−95.6) and a specificity of 26.5% (95% CI: 21.1−28.9).

**Table 2:** Sensitivity and specificity across multiple IP-10 cut-off values for detection of unsuppressed HIV VL (>1000 copies/mL), both overall and stratified by clinic type (stable, specialized risk).

| Cut-off<br>value | Overall (n=1051)<sup>a</sup> |  | Stable clinic (n=456) |  | Specialized risk clinic<br>(n=595) |  |
| --- | --- | --- | --- | --- | --- | --- |
|  | Sens (%) | Spec (%) | Sens (%) | Spec (%) | Sens (%) | Spec (%) |
| 6.4 | **92.91** | **26.84** | **91.67** | **37.79** | **92.86** | **16.98** |
| 6.5 | **92.13** | **28.14** | **91.67** | **39.40** | **91.96** | **18.01** |
| 6.6 | **91.34** | **30.30** | **91.67** | **41.47** | **91.07** | **20.29** |
| 6.7 | **91.34** | **32.36** | **91.67** | **44.47** | **91.07** | **21.53** |
| 6.8 | **90.55** | **33.77** | 83.33 | 45.39 | **91.07** | **23.40** |
| 6.9 | **90.55** | **35.28** | 83.33 | 47.00 | **91.07** | **24.84** |
| 7.0 | 89.76 | 36.58 | 83.33 | 47.93 | **90.18** | **26.50** |
| 7.1 | 88.96 | 38.20 | 83.33 | 50.00 | 89.29 | 27.54 |
| 7.2 | 85.83 | 39.61 | 75.00 | 52.07 | 86.61 | 28.36 |
| 7.3 | 83.46 | 41.45 | 75.00 | 53.97 | 83.93 | 30.23 |

Optimal thresholds for screening unsuppressed viral load were selected using a predefined criterion that prioritised the first threshold with sensitivity above 90%, as the IP-10 POC test was validated as a triage test. Accordingly, values highlighted in bold in the table indicate sensitivity above 90%.
<sup>a</sup>Six IP-10 POC test samples were excluded from this analysis because their corresponding VL results were not available
**Abbreviations:** Sens: sensitivity, Spec: specificity

Given the higher proportion of participants with LLV in specialized risk clinics compared to stable patient clinics, we assessed the impact of LLV on IP-10 classification outcomes by stratifying results by level of viremia (Table 3). Overall, 77.9% (n=74) of the 95 participants with LLV had a false positive result and would therefore be classified as having unsuppressed VL. When stratified by clinic type, in stable patient clinics, of the few LLV, 41% (n=7) had a false positive result. In specialized risk clinics 85.9% (n=67) had a false positive result.

**Table 3:** Clinical performance of the IP-10 point-of-care triage test for the detection of unsuppressed HIV viral load (VL) in terms of false positives, false negatives, true positives, true negatives stratified by clinic type (stable, specialized risk) and VL category (undetectable, low-level viremia, unsuppressed)^a^.

| Overall |  |  |  |  | Stable clinic |  |  |  | Specialized risk clinic |  |  |  |
| --- | --- | --- | --- | --- | --- | --- | --- | --- | --- | --- | --- | --- |
|  | Total<br>n=<br>1051<sup>b</sup> | Undetectable<br>n=829 | Low level<br>viremia<br>n=95 | Unsuppressed<br>n=127 | Total<br>n=<br>446<sup>c</sup> | Undetectable<br>n=417 | Low level<br>viremia<br>n=17 | Unsuppressed<br>n=12 | Total<br>n=<br>595<sup>c</sup> | Undetectable<br>n=405 | Low level<br>viremia)<br>n=78<sup>c</sup> | Unsuppressed<br>n=112<sup>c</sup> |
| **TP n (%)** | 116<br>(11.04) | NA | NA | 116 (91.34) | 11<br>(2.47) | NA | NA | 11 (91.67) | 102<br>(17.14) | NA | NA | 102 (91.07) |
| **FP n (%)** | 625<br>(59.47) | 551 (66.47) | 74 (77.89) | NA | 241<br>(54.04) | 234 (56.12) | 7 (41.18) | NA | 379<br>(63.70) | 312 (77.04) | 67 (85.90) | NA |
| **TN n (%)** | 299<br>(28.45) | 278 (33.53) | 21 (22.11) | NA | 193<br>(43.27) | 183 (43.88) | 10 (58.82) | NA | 104<br>(17.48) | 93 (22.96) | 11 (14.10) | NA |
| **FN n (%)** | 11<br>(1.05) | NA | NA | 11 (8.66) | 1<br>(0.22) | NA | NA | 1 (8.33) | 10<br>(1.68) | NA | NA | 10 (8.93) |

<sup>a</sup>Undetectable: (<50 copies/mL), LLV: Low level viremia (50-1000 copies/mL), Unsuppressed: (>1000 copies/mL)
<sup>b</sup>Six IP-10 POC test samples were excluded from this analysis because their corresponding VL results were not available
<sup>c</sup>Ten participants from other consultations are not shown in the table when stratifying by clinic type
**Abbreviations:** FP: false positives, FN: false negatives, NA: not applicable, TP: true positives, TN : true negatives

### Predictors of false positivity

Multivariable logistic regression revealed that the factors most strongly and independently associated with false positivity of the IP-10 POC test were obesity (adjusted odds ratio (aOR)= 3.47, 95% CI: 1.74−6.93), enrolment in a one-stop TB/HIV clinic (aOR=2.99, 95% CI: 1.09−8.15), and cotrimoxazole use in the previous year (aOR= 2.16, 95% CI: 1.13−4.13) (Table 4). Bivariable analysis results are shown in Table S1. One-stop TB/HIV clinic attendance and cotrimoxazole use in the previous year were more prevalent among individuals from specialized risk clinics compared with those from stable patient clinics (p <0.001) (Table 1, Table S1). Obesity was not associated with specialized risk clinics (p= 0.581).

**Table 4:**
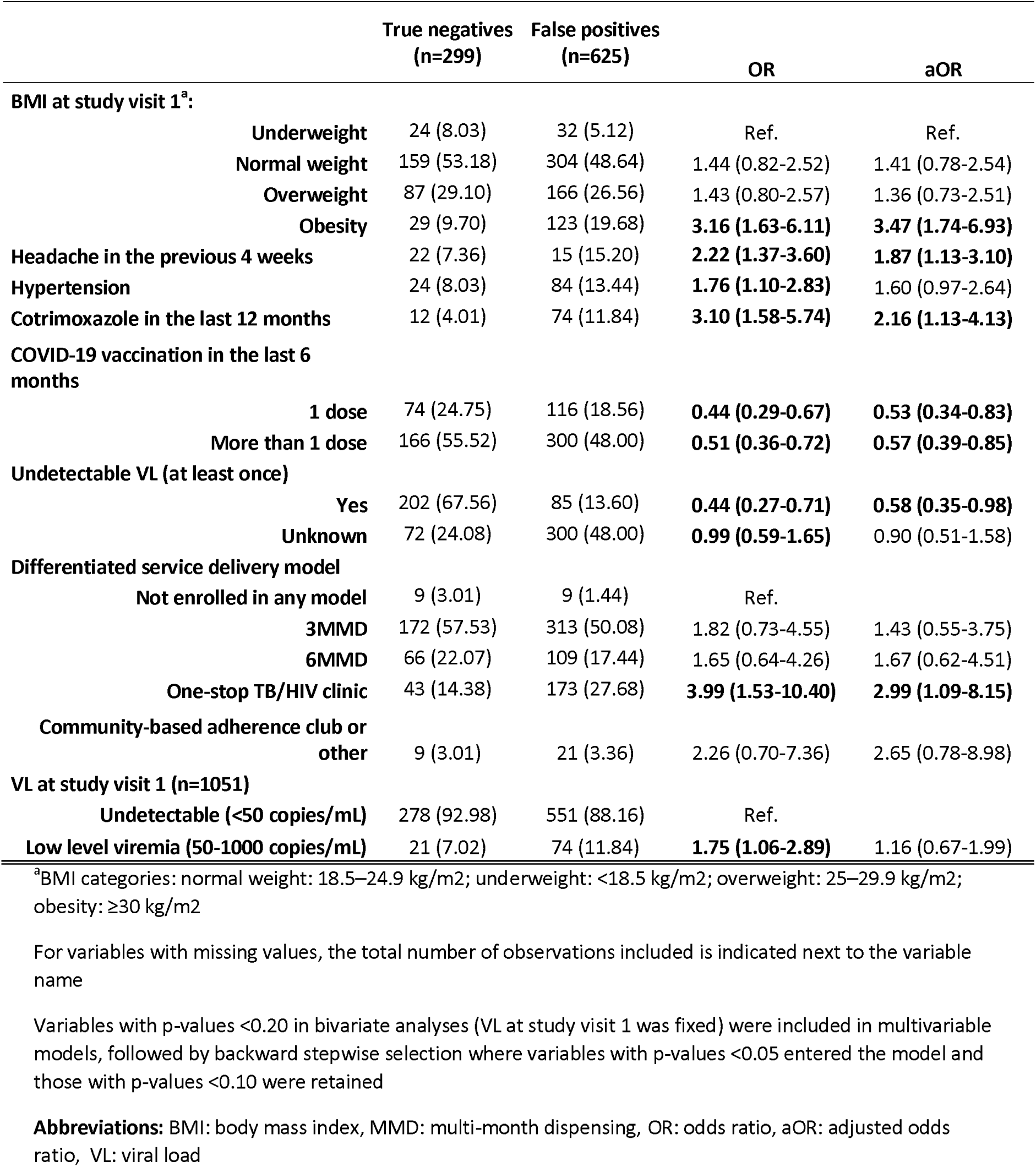
Multivariable analysis of factors associated with IP-10 point-of-care test false positivity. Analysis includes all individuals with an HIV viral load <1000 copies/mL (n=924). All values are presented as n (%).

On the other hand, receiving a COVID-19 vaccine in the last six months was associated with a lower risk of false positivity with aOR of 0.53 (95% CI: 0.34–0.83) for one dose (Table 1, Table S1). Additionally, having had at least one undetectable VL was associated with lower false positivity (aOR = 0.58, 95% CI: 0.35–0.98).

### Immediate end-user acceptability

Forty-three PLHIV on ART participated in exit interviews. The sample included more women (65.1%, n=28) than men, and more participants aged 26-49 years (72.1%, n=31) (Table 5). This distribution aligns with the expected age and gender profile of patients attending participating health facilities. Forty-two percent of participants (n=18) were long-term ART users (6-18 years on treatment), and 41.9% (n=18) reported variable treatment adherence over the previous three months. Seventy-four per cent (n=32) received a positive IP-10 POC test result based on the optimal threshold test reading value of 6.7 (Table 6).

**Table 5.**
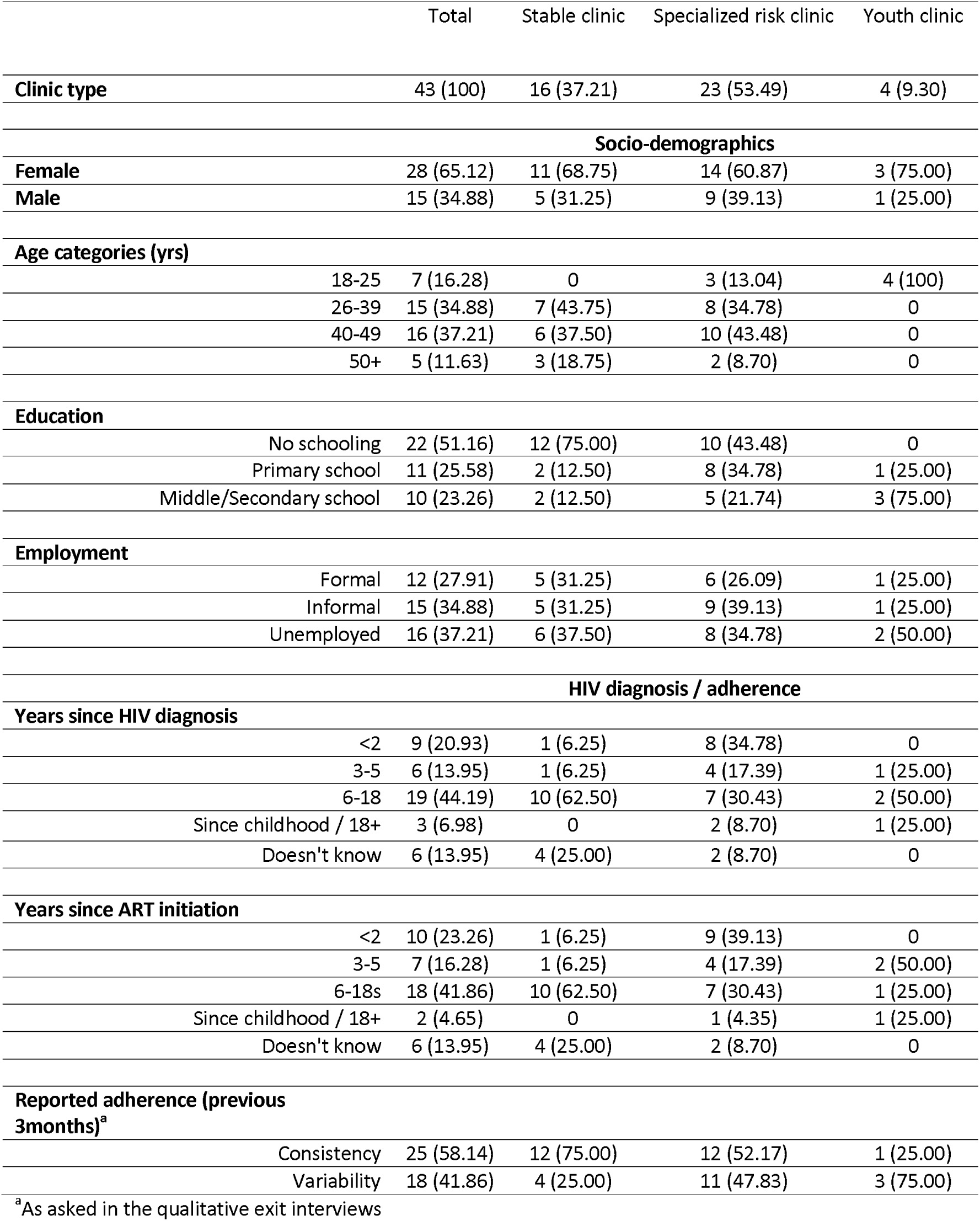
Characteristics of sampled participants for IP-10 POC test exit interviews, by clinic type (stable, specialized risk, youth). All values are presented as n (%).

|  | Total | Stable clinic | Specialized risk clinic | Youth clinic |
| --- | --- | --- | --- | --- |
| **Clinic type** | 43 (100) | 16 (37.21) | 23 (53.49) | 4 (9.30) |
| **Socio-demographics** |  |  |  |  |
| **Female** | 28 (65.12) | 11 (68.75) | 14 (60.87) | 3 (75.00) |
| **Male** | 15 (34.88) | 5 (31.25) | 9 (39.13) | 1 (25.00) |
| **Age categories (yrs)** |  |  |  |  |
| 18-25 | 7 (16.28) | 0 | 3 (13.04) | 4 (100) |
| 26-39 | 15 (34.88) | 7 (43.75) | 8 (34.78) | 0 |
| 40-49 | 16 (37.21) | 6 (37.50) | 10 (43.48) | 0 |
| 50+ | 5 (11.63) | 3 (18.75) | 2 (8.70) | 0 |
| **Education** |  |  |  |  |
| No schooling | 22 (51.16) | 12 (75.00) | 10 (43.48) | 0 |
| Primary school | 11 (25.58) | 2 (12.50) | 8 (34.78) | 1 (25.00) |
| Middle/Secondary school | 10 (23.26) | 2 (12.50) | 5 (21.74) | 3 (75.00) |
| **Employment** |  |  |  |  |
| Formal | 12 (27.91) | 5 (31.25) | 6 (26.09) | 1 (25.00) |
| Informal | 15 (34.88) | 5 (31.25) | 9 (39.13) | 1 (25.00) |
| Unemployed | 16 (37.21) | 6 (37.50) | 8 (34.78) | 2 (50.00) |
| **HIV diagnosis / adherence** |  |  |  |  |
| **Years since HIV diagnosis** |  |  |  |  |
| <2 | 9 (20.93) | 1 (6.25) | 8 (34.78) | 0 |
| 3-5 | 6 (13.95) | 1 (6.25) | 4 (17.39) | 1 (25.00) |
| 6-18 | 19 (44.19) | 10 (62.50) | 7 (30.43) | 2 (50.00) |
| Since childhood / 18+ | 3 (6.98) | 0 | 2 (8.70) | 1 (25.00) |
| Doesn't know | 6 (13.95) | 4 (25.00) | 2 (8.70) | 0 |
| **Years since ART initiation** |  |  |  |  |
| <2 | 10 (23.26) | 1 (6.25) | 9 (39.13) | 0 |
| 3-5 | 7 (16.28) | 1 (6.25) | 4 (17.39) | 2 (50.00) |
| 6-18s | 18 (41.86) | 10 (62.50) | 7 (30.43) | 1 (25.00) |
| Since childhood / 18+ | 2 (4.65) | 0 | 1 (4.35) | 1 (25.00) |
| Doesn't know | 6 (13.95) | 4 (25.00) | 2 (8.70) | 0 |
| **Reported adherence (previous 3months)<sup>a</sup>** |  |  |  |  |
| Consistency | 25 (58.14) | 12 (75.00) | 12 (52.17) | 1 (25.00) |
| Variability | 18 (41.86) | 4 (25.00) | 11 (47.83) | 3 (75.00) |

<sup>a</sup>As asked in the qualitative exit interviews

**Table 6.**
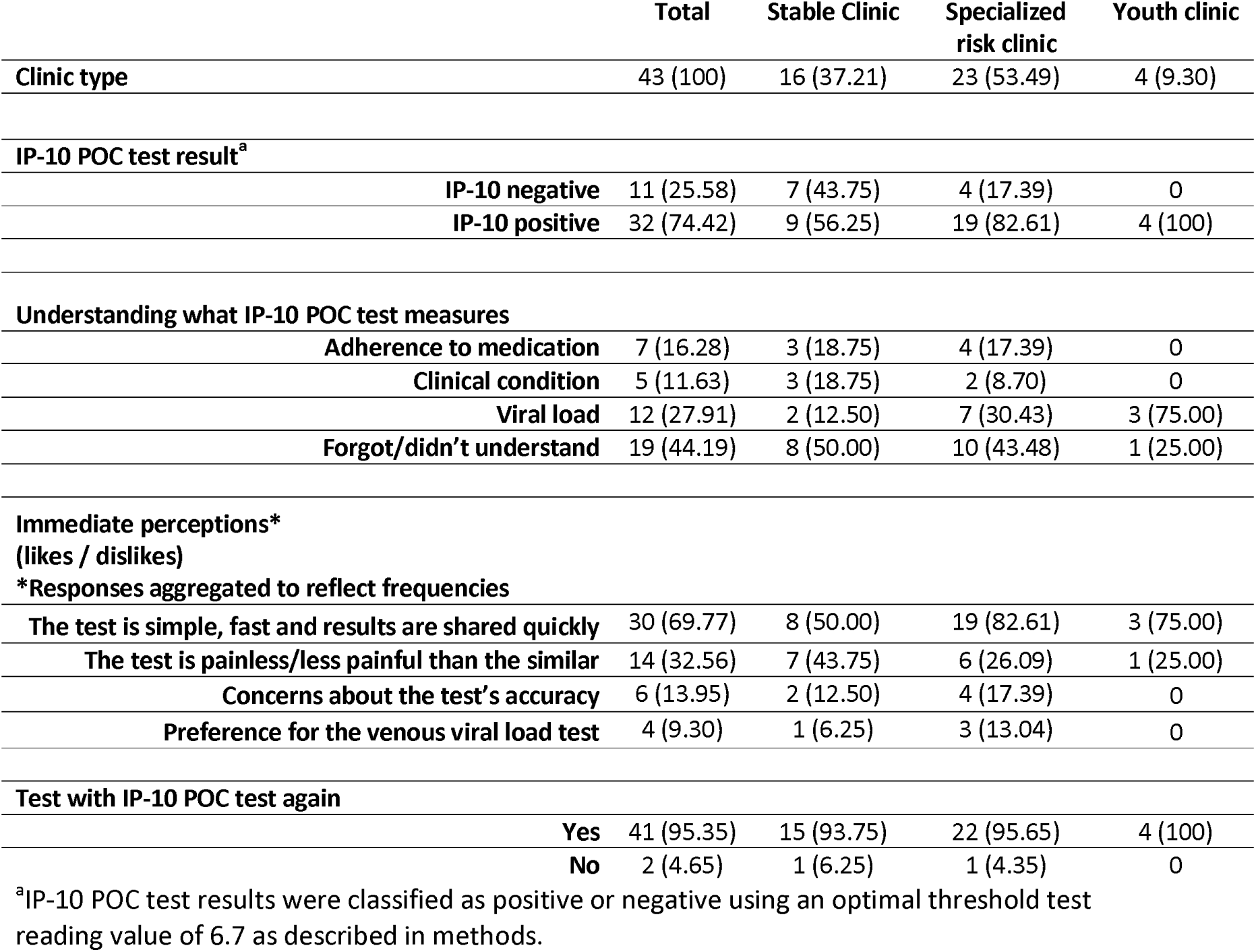
Perceptions and experiences of the IP-10 POC triage test, by clinic type (stable, specialized risk, youth). All values are presented as n (%).

|  | Total | Stable Clinic | Specialized risk clinic | Youth clinic |
| --- | --- | --- | --- | --- |
| **Clinic type** | 43 (100) | 16 (37.21) | 23 (53.49) | 4 (9.30) |
| **IP-10 POC test result<sup>a</sup>** |  |  |  |  |
| **IP-10 negative** | 11 (25.58) | 7 (43.75) | 4 (17.39) | 0 |
| **IP-10 positive** | 32 (74.42) | 9 (56.25) | 19 (82.61) | 4 (100) |
| **Understanding what IP-10 POC test measures** |  |  |  |  |
| **Adherence to medication** | 7 (16.28) | 3 (18.75) | 4 (17.39) | 0 |
| **Clinical condition** | 5 (11.63) | 3 (18.75) | 2 (8.70) | 0 |
| **Viral load** | 12 (27.91) | 2 (12.50) | 7 (30.43) | 3 (75.00) |
| **Forgot/didn't understand** | 19 (44.19) | 8 (50.00) | 10 (43.48) | 1 (25.00) |
| **Immediate perceptions\***<br>**(likes / dislikes)** |  |  |  |  |
| **\*Responses aggregated to reflect frequencies** |  |  |  |  |
| **The test is simple, fast and results are shared quickly** | 30 (69.77) | 8 (50.00) | 19 (82.61) | 3 (75.00) |
| **The test is painless/less painful than the similar** | 14 (32.56) | 7 (43.75) | 6 (26.09) | 1 (25.00) |
| **Concerns about the test's accuracy** | 6 (13.95) | 2 (12.50) | 4 (17.39) | 0 |
| **Preference for the venous viral load test** | 4 (9.30) | 1 (6.25) | 3 (13.04) | 0 |
| **Test with IP-10 POC test again** |  |  |  |  |
| **Yes** | 41 (95.35) | 15 (93.75) | 22 (95.65) | 4 (100) |
| **No** | 2 (4.65) | 1 (6.25) | 1 (4.35) | 0 |

<sup>a</sup>IP-10 POC test results were classified as positive or negative using an optimal threshold test reading value of 6.7 as described in methods.

Seventy percent (n=30) appreciated the test’s simplicity and the quick turnaround of results, with this view more common among participants in the specialized risk clinic (82.6%, n=19) vs the stable patient clinic (50.0%, n=8) (Table 6). Thirty-three percent (n=14) shared that the test was less painful and less invasive than the venous blood draw. A smaller proportion expressed concern about its accuracy (14.0%, n=6), citing unfamiliarity with the test, and 9.3% (n=4) shared their trust in the conventional laboratory-based VL test process, which they shared was lost with a POC finger prick test.

Understanding what the POC test measured was limited: participants described it as reflecting ART adherence (16.3%, n=7), clinical condition (11.6%, n=5), or VL (27.9%, n=12); nearly half (44.2%, n=19) were unsure or could not recall (Table 6). Despite this uncertainty, almost all participants (95.3%, n=41) expressed interest in being tested with the IP-10 POC test again. Perceptions and experiences of the test were broadly similar between the stable, specialized risk, and youth clinics.

## Discussion

This real-world clinical performance study demonstrated the operational feasibility of the IP-10 POC test for screening unsuppressed HIV VL. The study identified predictors of false-positive results, including cotrimoxazole use, enrolment in a one-stop TB/HIV clinic, and obesity. All the factors except for obesity were more prevalent among PLHIV attending specialized risk clinics. The results confirmed findings from South Africa, demonstrating that the IP-10 POC triage test performed as a highly sensitive, moderately specific triage tool. Most PLHIV welcomed the test as a less invasive immediate health indicator but the data also highlighted a limited understanding of the test’s purpose and concerns regarding the test’s reliability compared to laboratory-based VL testing.

A key finding was the decreased test specificity (26.5%) among participants attending specialized risk clinics catering to PLHIV with histories of detectable VL and/or adherence issues. These clinics had higher prevalences of ART interruptions, unsuppressed VL and LLV than did stable patient clinics, likely leading to lingering elevated levels of IP-10. In stable patient clinics, the performance of the IP-10 POC was similar to that of the South African study, with specificities of 44.5% and 43.6% in the Mozambique and South African studies, respectively. The South African study was conducted in stable PLHIV who had been on ART for at least 12 months without treatment interruptions in the previous three months (14).

To ascertain the best approach for optimizing the IP-10 POC test as a triage tool, we assessed risk factors associated with false positivity of the IP-10 POC test. Obesity, enrolment in a one-stop TB/HIV clinic (a DSD service for PLHIV co-infected and/or in treatment for TB), and cotrimoxazole use in the previous year (prescribed in immunocompromised patients with CD4 < 200 cells/mm^3^ for treatment or prevention of opportunistic infections) were independently associated with false positivity for the IP-10 POC test. These are likely proxies for systemic inflammation and/or IP-10 elevation. Plasma IP-10 levels have been reported to increase in response to various infections (SARS-CoV-2, tuberculosis, hepatitis C), as well as in some non-communicable diseases (18–25). Obesity has also been shown to trigger inflammatory processes including increases in adipose-tissue IP-10 levels (26). Indeed, IP-10 is a proinflammatory chemokine not specific for HIV infection. There are likely to be other non-measured factors potentially contributing to false positivity.

Our results revealed challenges in classification of PLHIV with LLV. While the IP-10 POC test uses a dichotomous VL 1,000 copies/mL threshold and should categorize LLV as suppressed, 40% were classified as unsuppressed, likely due to intermediate levels of IP-10. Although technically “false positives” for the IP-10 POC test (Table 3), this classification acts as a conservative approach that prioritizes sensitivity and would triage these patients toward WHO-recommended EAC and VL testing (11,27). Thus, using the IP-10 POC in stable patient clinics where LLV prevalence is low (<4%) would effectively limit the number of LLV who might miss the necessary EAC intervention.

DSD models are central to ART delivery across sub-Saharan Africa, including Mozambique, with most modalities designed for stable PLHIV, defined as individuals on ART for at least six months with at least one suppressed VL result in the past 6 months. DSD models primarily include community-based adherence groups, decentralized ART pick-up points, and multi-month (MMD) ART dispensing (28). In our study, as expected, stable patient clinic attendees were much more likely than specialized risk clinic attendees to be enrolled in six-month MMD (40% vs 0.7%) and showed superior VL suppression (93% vs 68%). Thus, the combination of the high likelihood of viral suppression and the superior performance of the IP-10 POC test positions stable PLHIV as the most appropriate target for IP-10 triage test implementation. Given these advantages, the current practice of universal individual VL testing among stable PLHIV may represent an inefficient use of limited and expensive laboratory resources. Combined with evidence of the test’s cost-effectiveness (14), our current findings suggest that integrating the IP-10 POC triage test into DSD stable care pathways, particularly within the 6MMD model could substantially reduce overall VL testing needs and enable health systems to reallocate VL testing capacity toward PLHIV requiring closer clinical attention. Potential integration pathways include using a positive IP-10 POC triage result as a gateway either to EAC, or to immediate laboratory-based or POC VL confirmation. In Mozambique, in 2025, an estimated 86% of the 1,947,886 PLHIV on ART were considered stable (28) representing over 1.6 million PLHIV who could benefit from such an approach. Acceptability data supports this strategy; patients preferred the finger-prick over venous draws, highlighting its resemblance to familiar HIV and malaria rapid tests. Concerns expressed regarding result reliability emphasize the importance of patient-centred communication regarding the purpose, integration into care and limitations of triage testing.

A significant limitation of the study is that it was designed to assess performance within the whole study population. It was not powered to assess performance of the IP-10 POC triage test stratified by clinic type and this resulted in imbalanced sample sizes, particularly for cases of unsuppressed VL with the majority coming from the specialized risk clinics. However, the optimal IP-10 POC test reading threshold was lower in the stable population as compared to the overall and is thus not likely to have led to overestimation of test specificity.

The study exhibits several strengths, notably the inclusion of end-user acceptability data alongside performance results. Additionally, performance metrics are more robust than the South African study due to a larger sample size. Furthermore, recruitment from specialized risk clinics allowed for the identification of actionable clinical factors associated with false positivity which will be crucial for refining the IP-10 POC triage test’s use case.

## Conclusions

While universal VL testing remains the gold-standard, in certain settings, introduction of a triage test offers an alternative for optimizing the balance between VL availability, health system costs, provision of routine care for stable PLHIV, and preservation of resources for PLHIV in need of specialized support. This is particularly important in recent times of significant reduction of international funding for fighting HIV in low resource settings.

## Authors’ contributions

DN, ASL, DAB and HM designed the research study. DAB, MFC, ASL and EB, collected and validated the data. DN, ASL and EB contributed to clinical data analysis. HM and LV performed the qualitative analysis of exit interviews. DN drafted the manuscript. ASL, TRW, EB, HM and PV performed data review and critical revision of the manuscript. All authors reviewed, gave critical input and approved the final version of the manuscript.

## Supporting information

Saura_IP_10_supplementary methods

Saura_IP_10_supplementary Table

## Acknowledgements

The authors would like to thank the study participants and communities for their invaluable contribution to this work. We acknowledge support from the grant CEX2023-0001290-S funded by MCIN/AEI/ 10.13039/501100011033, and support from the Generalitat de Catalunya through the CERCA Program We acknowledge the support of the of the Ministry of Health of Mozambique, Maputo Province health directorate and Matola and Magude district health services (Serviços Distritais de Saúde, Mulher e Acção Social – SDSMAS) for their collaboration in implementation. We are grateful to the study nurses Nelly Micael and Balbina Cumba, Health Facility medical officer Cristina Silindane, laboratory technicians Paulina Macamo and Cristeza Samuel, for their assistance with patient recruitment and data collection.

## Data availability statement

The anonymised data that support the findings of this study are available from the corresponding author upon reasonable request.

## Supporting Information

**Appendix 1: Supplementary methods** 1) for the Magude and Matola Health facilities participating in the study methods and 2) for the qualitative study including the discussion guide for exit interviews.

**Appendix 2: Supplementary results** Table S1: Bivariable and multivariable analysis of factors associated with IP-10 point-of-care test false positivity.

