## Supplementary material for "Determinants of specificity and end-user acceptability of an IP-10-based point-of-care triage test for antiretroviral therapy monitoring in Mozambique": Saura_IP_10_supplementary methods: Saura_IP_10_supplementary methods.docx

**APPENDIX I: SUPPLEMENTARY METHODS**

**Supplementary methods for the Magude and Matola Health facilities participating in the study**

Magude is a rural district located in the northern part of Maputo province, bordering South Africa. One of its nine health facilities was included in this study as a high-volume facility with 5,211 active individuals on ART. Matola is an urban district near Maputo city and we included one of the 26 facilities in this study as a high-volume facility with more than 1,000 clients on ART. HIV treatment and care are provided free of charge, with VL testing recommended annually for ART monitoring (1).

Stable patients—defined as those on ART with sustained viral suppression and good adherence—are managed through extended clinical follow-up intervals and multi-month dispensing. In contrast, patients classified as at-risk receive more intensive clinical and laboratory monitoring, including frequent consultations and enhanced adherence support, to address potential treatment failure and improve outcomes. This dual approach allows the health system to maintain high-quality, patient-centred care while improving efficiency and focusing clinical attention on those most in need.

Youth clinics are a one-stop service for adolescents and young people up to 24 years of age offering counselling, testing and treatment for HIV and other sexually transmitted infections and were recruited for the qualitative acceptability study.

**REFERENCE:**

1. Guião de cuidados do HIV do Adulto, Adolescente, Grávida, lactante e criança. Ministério da Saúde de Moçambique, 2023.

**Supplementary methods for the qualitative study including the discussion guide for exit interviews.**

**Title of the research project: Piloting a novel antiretroviral therapy monitoring algorithm with a new rapid screening test for HIV virological failure in Mozambique (VIP 2 study)**

**Discussion guide** for patients, round 1 exit interview

**Purpose of the guide:** VIP 2 is a project to evaluate the clinical performance of a point-of-care IP-10 point of care (POC) test for screening for virological failure in ART-treated people living with HIV (PLHIV) using finger prick blood specimens. The project includes evaluation of the acceptability and feasibility of implementing the point-of-care IP-10 POC into Mozambique’s routine guidelines for ART monitoring. The acceptability and feasibility component includes between one and three interviews with participants enrolled in the clinical performance component of the project at three different study time points.

This discussion guide is to be used with participants as a rapid exit interview following their IP-10 POC testing and result. The guide covers 4 key topic areas to briefly explore participant background, treatment adherence, HIV/ART monitoring, and their immediate perceptions and experiences of the IP-10 POC test.

**Form of data recording:** (1) Audio recording (2) Key points handwritten next to each question in the space provided (3) Capturing each participant’s answers into the relevant study template.

**Expected time needed per use:** 10-20 minutes

**Instructions***:* Use the below questions and prompts to engage the participant in conversation after following the informed consent process.

Preamble (**to be read by research assistant**): Today is the (**insert date [day xx^th^ Xxx xxxx**]) and it is (**insert time XX:XX**). This is a discussion with a patient enrolled in the VIP2 study at (**insert clinic name**). Thank you for your time. These discussions are part of the VIP 2 study which aims to understand the experiences of patients about viral load monitoring and IP-10 POC test. This discussion will cover 4 topic areas: questions about your personal background, treatment adherence, monitoring, and your experience of the IP-10 point-of-care test.

I want to remind you that there are no right or wrong answers — you’re the expert of your life and experiences, and I am happy to be learning from you. Do you have any questions before we begin?

| **Topic** | **Questions/probes** |
| --- | --- |
| **Personal**  **details** | 1. Age: ________ 2. Male / female: ______________________________________ 3. Education/schooling completed: _____________________________________ 4. Employment: ____________________________________________________ 5. Religion: ________________________________________________________ 6. Home language(s): _______________________________________________ |
| **Treatment adherence** | 1. What year were you diagnosed with HIV? 2. What year did you first start treatment for HIV (ART)? 3. How long have you been on ART? 4. Research shows that for many people, adherence to treatment goes up and down over time. What is your experience of adhering to ART? 5. How would you describe your adherence over the last three months (probe for disruptions, challenges, support received)? |
| **Viral load monitoring** | 1. Please tell me what you understand with the term ‘viral load’? 2. Where did you hear about viral load the first time? How was ‘viral load’ explained to you? 3. How often do you have a viral load test done at the clinic? How are the results shared with you? 4. Is the viral load test a useful test to you? Why or why not? 5. How do you think adherence affects viral load? |
| **Perceptions, experiences of the point-of-care IP-10** | 1. As part of this study, you just had the IP-10 finger prick test. Tell me, how do you understand what the test is for? 2. What was the test result? How did you understand what the results meant? 3. Did the results surprise you (why or why not)? Did you have any questions? What were they, if so? 4. Likes/dislikes. What did you like about the IP-10 test? What did you dislike? 5. How did the finger prick viral load test compare to other health tests you’ve done? 6. What are the benefits / value of having a finger prick test potentially show if your viral load is high or low? 7. What do you think are some disadvantages/potential problems with this test? 8. Would you have this test again if it was offered? Why? |
| **Closing** | Summarise the key points for respondent checking.  Do you have any questions or points to raise? |
| Thank you for your time today, we appreciate your participation in the study and for sharing openly about your experiences. As explained to you during the consent process, you will receive a telephone call from a study nurse with the results of your serum/blood viral load test in about 15 days.  Because the finger prick test is not 100 accurate, these results could be different to the results of your finger prick test (for example, the viral load blood test could show positive for a high viral load while your finger prick test showed negative, or vice versa). We are interested in having follow-up interviews with people as they receive these results to include the experiences of different patients.  These interviews will be a bit longer (between 30-45 mins) and will ask about your experiences of living with HIV, adhering to treatment, HIV monitoring, and the different tests in more detail. Would you like to continue to be part of these interviews? If so, please share your contact details so I may get in touch with you. Your details will not be used for any other purpose. Thank you again. | |
