## Supplementary material for "Determinants of specificity and end-user acceptability of an IP-10-based point-of-care triage test for antiretroviral therapy monitoring in Mozambique": Saura_IP_10_supplementary Table: Saura_IP_10_supplementary Table.docx

**APPENDIX II: SUPPLEMENTARY RESULTS**

**Table S1:** Bivariable and multivariable analysis of factors associated with IP-10 point-of-care test false positivity. Analysis includes all individuals with an HIV viral load <1000 copies/mL (n=924). All values are presented as n (%).

|  | **True negatives (n=299)** | **False positives (n=625)** | **OR^a^** | **aOR^b^** |
| --- | --- | --- | --- | --- |
| **Socio-demographics** | | | | |
| **Biological sex (female)** | 204 (68.23) | 456 (72.96) | 1.26 (0.93-1.70) |  |
| **Age (years)^c^** | 41.73 (36.16-48.61) | 41.89 (34.82-49.42) | 1.00 (0.99-1.01) |  |
| **Employment** |  |  |  |  |
| **Employed** | 91 (30.43) | 224 (35.84) | Ref. |  |
| **Part-time** | 78 (26.09) | 140 (22.40) | 0.73 (0.50-1.05) |  |
| **Not employed** | 130 (43.48) | 261 (41.76) | 0.82 (0.59-1.12) |  |
| **Marital status** |  |  |  |  |
| **Married^e^** | 156 (52.17) | 336 (53.60) | Ref. |  |
| **Single** | 110 (36.79) | 211 (33.76) | 0.89 (0.66-1.20) |  |
| **Divorced** | 2 (0.67) | 6 (0.96) | 1.21 (0.28-5.28) |  |
| **Widowed** | 31 (10.37) | 73 (11.68) | 1.09 (0.69-1.72) |  |
| **Educational level** |  |  |  |  |
| **No primary school** | 52 (17.30) | 97 (15.52) | Ref. |  |
| **Primary school** | 165 (55.18) | 315 (50.40) | 1.03 (0.70-1.51) |  |
| **Secondary school** | 75 (25.08) | 197 (31.52) | 1.41 (0.92-2.16) |  |
| **Post-secondary school or university** | 7 (2.34) | 16 (2.564) | 1.18 (0.45-2.99) |  |
| **Signs and symptoms at study visit 1** | | | | |
| **Clinic** |  |  |  |  |
| **Stable** | 193 (64.55) | 241 (38.56) | Ref. |  |
| **Specialized risk** | 104 (34.78) | 379 (60.64) | **2.91 (2.18-3.88**)^d^ |  |
| **Other** | 2 (0.67) | 5 (0.80) | 1.77 (0.39-7.95) |  |
| **BMI^f^** |  |  |  |  |
| **Underweight** | 24 (8.03) | 32 (5.12) | Ref. | Ref. |
| **Normal weight** | 159 (53.18) | 304 (48.64) | 1.44 (0.82-2.52) | 1.41 (0.78-2.54) |
| **Overweight** | 87 (29.10) | 166 (26.56) | 1.43 (0.80-2.57) | 1.36 (0.73-2.51) |
| **Obesity** | 29 (9.70) | 123 (19.68) | **3.16 (1.63-6.11)** | **3.47 (1.74-6.93)** |
| **Temperature (Celsius)^c^** | 36.2 (36.0-36.4) | 36.2 (36.0-36.4) | 1.03 (0.87-1.23) |  |
| **Blood pressure (n=876)^g^** |  |  |  |  |
| **Normal** | 99 (35.11) | 178 (29.97) | Ref. |  |
| **Elevated** | 183 (64.89) | 416 (70.03) | **1.27 (0.94-1.71)** |  |
| **Cough** |  |  |  |  |
| **No** | 289 (96.66) | 582 (93.12) | Ref. |  |
| **Yes (cough for 2 weeks or less)** | 6 (2.01) | 10 (1.60) | 0.80 (0.30-2.16) |  |
| **Yes (cough for over 2 weeks)** | 4 (1.34) | 33 (5.28) | **3.70 (1.37-10.00)** |  |
| **Sore throat** | 7 (2.34) | 20 (3.20) | 1.32 (0.57-3.08) |  |
| **Shortness of breath or difficulty breathing** | 3 (1.00) | 9 (1.44) | 1.31 (0.38-4.48) |  |
| **Muscle pain** | 16 (5.35) | 36 (5.76) | 1.06 (0.58-1.94) |  |
| **Tiredness** | 11 (3.68) | 30 (4.80) | 1.28 (0.64-2.57) |  |
| **Headache** | 15 (5.02) | 47 (7.52) | 1.51 (0.83-2.72) |  |
| **Diarrhoea** | 0 | 5 (0.80) | 5.31 (0.28-96.33) |  |
| **Vomiting** | 1 (0.33) | 0 | 0.16 (0.01-3.92) |  |
| **Skin eruption** | 7 (2.34) | 10 (1.60) | 0.67 (0.26-1.72) |  |
| **Signs and symptoms in the previous 4 weeks** | | | | |
| **Fever** | 26 (8.70) | 79 (12.64) | **1.50 (0.94-2.39)** |  |
| **Sore throat** | 11 (3.69) | 29 (4.64) | 1.24 (0.62-2.48) |  |
| **Shortness of breath or difficulty breathing** | 9 (3.01) | 19 (3.04) | 0.98 (0.45-2.15) |  |
| **Muscle pain** | 15 (5.02) | 52 (8.32) | **1.68 (0.94-3.01)** |  |
| **Tiredness** | 21 (7.02) | 50 (8.00) | 1.14 (0.67-1.92) |  |
| **Headache** | 22 (7.36) | 15 (15.20) | **2.22 (1.37-3.60)** | **1.87 (1.13-3.10)** |
| **Diarrhoea** | 6 (2.01) | 18 (2.88) | 1.38 (0.56-3.40) |  |
| **Vomiting** | 4 (1.34) | 7 (1.12) | 0.80 (0.25-2.58) |  |
| **Skin eruption** | 8 (2.68) | 24 (3.84) | 1.40 (0.63-3.09) |  |
| **Night sweats** | 5 (1.67) | 9 (1.44) | 0.83 (0.29-2.38) |  |
| **Comorbidities** | | | | |
| **Diabetes** | 0 | 1 (0.16) | 1.55 (0.06-38.12) |  |
| **Hypertension** | 24 (8.03) | 84 (13.44) | **1.76 (1.10-2.83)** | 1.60 (0.97-2.64) |
| **Epilepsy** | 1 (0.33) | 6 (0.96) | 2.09 (0.35-12.39) |  |
| **Past tuberculosis^h^** | 10 (3.34) | 32 (5.12) | 1.51 (0.74-3.06) |  |
| **Preventive tuberculosis treatment in the last 12 months (isoniazid)** | 51 (17.06) | 172 (27.52) | **1.84 (1.30-2.61)** |  |
| **Cotrimoxazole in the last 12 months** | 12 (4.01) | 74 (11.84) | **3.10 (1.58-5.74)** | **2.16 (1.13-4.13)** |
| **Previous confirmed COVID-19 in the last 3 months** | 1 (0.33) | 8 (1.28) | 2.73 (0.48-15.59) |  |
| **COVID-19 vaccination in the last 6 months** |  |  |  |  |
| **1 dose** | 74 (24.75) | 116 (18.56) | **0.44 (0.29-0.67)** | **0.53 (0.34-0.83)** |
| **More than 1 dose** | 166 (55.52) | 300 (48.00) | **0.51 (0.36-0.72)** | **0.57 (0.39-0.85)** |
| **Diagnosis or suspicion of malaria** | 5 (1.67) | 17 (2.72) | 1.53 (0.58-4.04) |  |
| **HIV clinical history** | | | | |
| **WHO stage at ART initiation** |  |  |  |  |
| **I or II** | 250 (83.61) | 533 (84.74) | Ref. |  |
| **III or IV** | 48 (16.05) | 138 (14.94) | 0.88 (0.60-1.28) |  |
| **Unknown** | 1 (0.33) | 2 (0.32) | 0.78 (0.10-5.96) |  |
| **WHO stage at study visit 1** |  |  |  |  |
| **I or II** | 245(81.94) | 516 (82.56) | Ref. |  |
| **III or IV** | 54 (18.06) | 109 (17.44) | 0.95 (0.67-1.37) |  |
| **ART regiment at ART initiation** |  |  |  |  |
| **DTG+2NRTI** | 133 (44.48) | 320 (51.20) | Ref. |  |
| **NNRTI+2NRTI** | 155 (51.84) | 295 (47.20) | 0.79 (0.60-1.05) |  |
| **Other** | 11 (3.68) | 10 (1.60) | 0.38 (0.16-0.90) |  |
| **Current ART regimen** |  |  |  |  |
| **DTG+2NRTI** | 294 (98.33) | 608 (97.28) | Ref. |  |
| **(NNRTI or PI) +2NRTI** | 5 (1.67) | 13 (2.08) | 1.19 (0.44-3.23) |  |
| **Other** | 0 | 4 (0.64) | 4.36 (0.23-81.17) |  |
| **Undetectable VL (at least once)** |  |  |  |  |
| **Yes** | 202 (67.56) | 85 (13.60) | **0.44 (0.27-0.71)** | **0.58 (0.35-0.98)** |
| **Unknown** | 72 (24.08) | 300 (48.00) | **0.99 (0.59-1.65)** | 0.90 (0.51-1.58) |
| **Differentiated service delivery model** |  |  |  |  |
| **Not enrolled in any model** | 9 (3.01) | 9 (1.44) | Ref. |  |
| **3MMD** | 172 (57.53) | 313 (50.08) | 1.82 (0.73-4.55) | 1.43 (0.55-3.75) |
| **6MMD** | 66 (22.07) | 109 (17.44) | 1.65 (0.64-4.26) | 1.67 (0.62-4.51) |
| **One-stop TB/HIV clinic** | 43 (14.38) | 173 (27.68) | **3.99 (1.53-10.40)** | **2.99 (1.09-8.15)** |
| **Community-based adherence club or other** | 9 (3.01) | 21 (3.36) | 2.26 (0.70-7.36) | 2.65 (0.78-8.98) |
| **Missed ART during the last month** |  |  |  |  |
| **None** | 232 (77.59) | 414 (66.24) | Ref. |  |
| **Any missed dose^i^** | 67 (22.41) | 211 (33.76) | **1.77 (1.28-2.41)** |  |
| **ART interruption (at least once) since ART initiation** | 18 (6.02) | 64 (10.24) | **1.75 (1.02-2.99)** |  |
| **VL at study visit 1** | | | | |
| **VL (n=1051)** |  |  |  |  |
| **Undetectable (<50 copies/mL)** | 278 (92.98) | 551 (88.16) | Ref. |  |
| **Low level viremia (50-1000 copies/mL)** | 21 (7.02) | 74 (11.84) | **1.75 (1.06-2.89)** | 1.16 (0.67-1.99) |

^a^Only variables with a p-value <0.20 in the bivariable analysis are highlighted in black

^b^Variables with p-values <0.20 in bivariate analyses (VL at study visit 1 was fixed) were included in multivariable models, followed by backward stepwise selection where variables with p-values <0.05 entered the model and those with p-values <0.10 were retained

^c^Median (Interquartile range)

^d^Clinic type was not included in the multivariable model because it cannot act as a predictive factor in clinical practice. Moreover, predictors retained in the final model differed statistically significantly between individuals attending the specialized risk clinic and those attending the stable clinic

^e^This category includes married participants, civil union or legal partnership

^f^BMI categories: normal weight: 18.5–24.9 kg/m2; underweight: <18.5 kg/m2; overweight: 25–29.9 kg/m2; obesity: ≥30 kg/m2

^g^Blood pressure categories: normal: systolic <120 mmHg and diastolic <80 mmHg; elevated: systolic 120-139 mmHg and/or diastolic 80-89 mmHg, or systolic 140-159 mmHg and/or diastolic 90-99 mmHg, or systolic 160-179 mmHg and/or diastolic 100-109 mmHg, or systolic ≥180 mmHg and/or diastolic ≥110 mmHg

^h^Tuberculosis site includes pulmonal, extrapulmonary, and unknown site

^i^This category includes at least one ART dose missed in the month

For variables with missing values, the total number of observations included is indicated next to the variable name

**Abbreviations**: ART: antiretroviral therapy, BMI: body mass index, DTG: dolutegravir, MMD: multi-month dispensing, NNRTI: non-nucleoside reverse transcriptase inhibitor, NRTI: nucleoside reverse transcriptase inhibitor, PI: Protease Inhibitor, POC: point-of-care, VL: viral load, WHO: World Health Organisation
